# Retrospective Analysis of HIV Pre-exposure Prophylaxis (PrEP) Awards Under Executive Order 14168 on Gender Ideology in the U.S. 2012-2025

**DOI:** 10.1101/2025.03.03.25323246

**Authors:** Evan Hall

**Affiliations:** School of Population and Public Health, University of British Columbia

**Author notes:** Correspondence Address: Evan Hall, School of Population and Public Health University of British Columbia 2206 East Mall Vancouver V6T 1Z3 Canada.

**Keywords:** HIV PrEP, policy, funding, healthcare, transgender, gender diverse people

## Abstract

**Background:** HIV PrEP-specific research is vital to advancing better health outcomes and reducing HIV transmissions in the United States. Executive Order (EO) 14168 targeting gender ideology will narrow the scope of research on gender and sex by research institutions. Implementation of EO 14168 is limiting communications and manuscript publications based on restricted terms, including “gender”, “transgender”, “LGBT” and more.

**Methods:** Award titles and abstracts were sourced from the Tracking Accountability in Government Grants System (TAGGS) for “PrEP” or “pre-exposure prophylaxis” relevant to HIV from 2012 to 2025. These award titles and abstracts were coded based on inclusion of restricted terms from EO 14168. The characteristics and amounts for award disbursements were analyzed. Data was subset based on elected presidential budgetary periods for political analysis.

**Results:** Of 388 unique award titles, 118 (30.4%) would be considered excluded based on the restricted terms of EO 14168. Transgender and gender were most represented restricted terminology. Mental health research grants (n = 244; 39.8%) compromised the majority of excluded award disbursements. Both Democrat and Republican states saw reductions in total HIV PrEP-specific research funding. The amount of excluded award disbursements totaled nearly $160 million (USD) from restricted terms.

**Discussion:** Applied retrospectively, EO 14168 and its implementation would have reduced the total funding to HIV PrEP-specific research by nearly $160 million (USD), impacting economic activity from the NIH by $400 million (USD) from 2012 to 2025.

**Highlights:**

- EO 14168 on gender ideology greatly impacts HIV PrEP-specific research
- $160 million (USD) lost if EO 14168 was applied retrospectively
- Gender and transgender most frequent restricted terms
- Mental health research grants most likely to be negatively impacted
- Nearly $400 million (USD) of economic activity potentially lost since 2012

## 1.0 Introduction

Human immunodeficiency virus (HIV) remains an ongoing epidemic in the United States, with 37,981 new HIV diagnoses reported in 2022 (1). HIV pre-exposure prophylaxis (PrEP) is a highly effective biomedical tool for preventing the transmission of HIV. The U.S. Food and Drug Administration (FDA) approved oral HIV PrEP in 2012 under its brand name (Truvada) (2). The FDA approved the release of the generic formula to market in 2020 (3). In 2022, the first long-acting injectable option for HIV PrEP was put to market, offering a bimonthly choice of administration (4). Yet, the brand price remains inaccessible to many individuals vulnerable to HIV in the United States (5). The USPSTF grade A rating of HIV PrEP has allowed for broader access to those on commercial or ACA marketplace insurance plans for free and at no cost, including lab and ancillary services (6). HIV PrEP has shown numerous high-impact outcomes in reducing the spread of HIV in the United States (7). However, under the current political climate and quick-changing shifts in research and health policy, the implementation and advancement of HIV PrEP are put into question.

The Trump Administration issued Executive Order (EO) 14168 targeting terminology that falls beyond the ‘gender’ binary and gender ideology (8). “Defending Women from gender ideology extremism and Restoring Biological Truth to the Federal Government” was an EO issued on January 20th, 2025, stating in part that “Federal funds shall not be used to promote gender ideology. Each agency shall assess grant conditions and grantee preferences and ensure grant funds do not promote gender ideology” (8). On the weekend of January 31st, 2025, the Centers for Disease Control and Prevention (CDC) began modifying website links, stating, “CDC’s website is being modified to comply with President Trump’s Executive Orders” (9). A similar process has occurred at the National Institutes of Health (NIH) (10). Beyond website content and communications, the CDC requested the withdrawal of manuscripts written or co-written by CDC scientists based on restricted terms referenced by EO 14168 (11). The restricted terms include gender, transgender, pregnant person, pregnant people, LGBT, transsexual, nonbinary, assigned male at birth, biologically male, and biologically female (12).

Gender, sexual orientation, and gender expression are distinct and non-interchangeable terms for specific social constructs (13). Additionally, sex can be categorized based on major components: chromosomal, gonadal, and phenotypic or anatomic, explaining how sex is not binary (14). These identities and categorizations, which are often intersectional, are closely related to HIV-related inequities in accessing HIV prevention, care, and treatment services (15–18). Outlined in the National HIV/AIDS Strategy from 2022-2025, priority populations included gay, bisexual, and other men who have sex with men and transgender women, stating that efforts are “essential if the nation is to succeed on the path toward the end the HIV epidemic by 2030” (19). The NIH Strategic Plan for HIV and HIV-Related Research (FY 2021-2025) also cites HIV PrEP-specific research as a crucial component to reducing new HIV transmissions (20). The Ending the HIV Epidemic (EHE) Initiative is an ambitious plan approved under the first Trump Administration in 2019 to prevent 255,000 new HIV cases by 2030 (21). From 2019 to 2023, Congress approved $1.8 billion ($USD) for EHE, including $774 million for CDC EHE activities (22). EHE projects are directly addressing the needs of sexual and gender minorities (23). EO 14168 will likely impact strategies, initiatives, and grant projects, including HIV PrEP, for HIV research and its implementation in the future. The prospective public health and financial outcomes are uncertain.

The ramifications of this Executive Order are challenging to describe based on various government agencies’ prospective implementation of these executive orders. However, it is clear that the HIV funding landscape, including for HIV PrEP, will be greatly altered in the coming years. Quantified measurements that characterize the detriments of these Executive Orders are necessary to understand the extent of the Order’s impact. This paper presents a retrospective analysis of how this executive Order shaped the HIV PrEP research landscape from 2012 to 2025. The results from this analysis offer a reference scenario to understand future changes in funding and themes in HIV PrEP-specific research.

## 2.0 Methods

Data was collected from the Tracking Accountability in Government Grants System (TAGGS) from the Department of Health and Human Services (HHS). The search criteria were specific to award titles from 2012 to 2025 that contained “pre-exposure prophylaxis” or “PrEP.” These award title entries were screened for relevance to HIV, where the researcher assessed contextualization of the award title topic. Additionally, abstracts were reviewed to confirm topical relevance to HIV. Abstracts were sourced from the NIH Research Portfolio Online Reporting Tools Expenditures and Results (RePORTER). Award titles and abstracts that were not context-specific to HIV were excluded from the dataset. These award disbursements comprised the “complete grant” dataset, including retained and excluded grants. The variables collected from TAGGS included: “Issue Date Fiscal Year,” (operational division) “OPDIV”, “Legal Entity Name”, “Legal Entity State”, “Legal Entity Congressional District”, “Legal Entity County” (unique entity identifier) “UEI”, “Recipient Type”, “Award Number,” “Award Title,” “Award Code,” “Budget Year,” “Award Class,” “Award Activity Type,” “Award Action Type” (assistance listing number) “ALN,” “Assistance Listing Title”; “Funding Fiscal Year”; “Sum of Actions.” Thes, the award number was cross-referenced to the award to identify unique awardees title, removing duplicates from which different institutions received different funding allotments from the same award title disbursement.

All abstracts from the complete dataset were sourced from RePORTER based on their award title and number. The list of terms implemented by EO 14168 is: “gender,” “transgender,” “pregnant person,” “pregnant people,” “LGBT,” “transsexual,” “nonbinary,” “assigned male at birth,” “biologically male,” and “biologically female,” which were sourced from leaked internal CDC communications (12). “LGBT” is an acronym for lesbian, gay, bisexual and trans, where “trans” was not included in this group, but all other terms were searched. Additionally, “nonbinary” was searched as “non-binary”. This paper refers to these terms as “restricted terms.” Each award title and its associated abstract were coded for each of the restricted terms, where “1” signified the term was present and “0” when the term was not. Award titles and abstracts may have had multiple restricted terms from the list. Then, the unique awards were identified based on whether or not they had any restricted terms containing “1” or only “0”. This output provides what this analysis categorizes as “excluded grants” (if any contained a restricted term), while grants that did not include any restricted terms are referred to as “retained grants.” An average across all of the possible excluded terms was taken. The unique award titles and abstracts were coded for a theoretical scenario if “men who have sex with men” (MSM) was included as a restricted term, where the same coding process applied, and a total number of unique awards and an average number of restricted terms were calculated. The primary dataset comprised award disbursements and not of unique award titles, as each award varied in length and may have been disbursed across multiple recipients. Two subsequent dataset of award disbursements was created, where (1) only containing retained grant data; (2) only containing excluded grant data. Award Activity Type, operational division, recipient type, and assistance listing title were counted, and percentages were calculated across the complete, retained, and excluded grant datasets based on award disbursements. The legal entity state entity data was converted into proportions based on the award disbursements analyzed from the complete, retained, and excluded grant datasets.

Political analysis was conducted on the complete, retained, and excluded grant datasets. The award disbursements were categorized based on elected presidential budgetary periods from 2012 to 2025, with four categories created “2012”, “2013-2017”, “2018-2021”, and “2022-2025” based on which political party held the presidency during the proposed presidential fiscal year. States from these award disbursements were categorized under the political party designation “Democrat” or “Republican” based on which political party received the majority of electoral college votes for that presidential election: 2008, 2012, 2016, and 2020 (24). Jurisdictions that did not participate in the presidential election, including Puerto Rico, Ontario, the Western Cape, and those unlabeled in the TAGGS, were categorized as “Other”.” The award amounts from the complete grant dataset were totaled for each elected presidential budgetary period for each party designation, with all award disbursements accounted for in each period, and were broken down by political party designation. Then, the retained grant dataset underwent the same data subsetting for award totals. The percentage of each political party designation (democrat, republican, other) was calculated in a bar plot. The total difference from award disbursement totals from the excluded grant dataset was calculated based on the totals of the complete grant dataset minus the retained grant dataset. Award disbursement amounts (in $USD) were derived from “Sum of Actions”. Based on the observational aim of this research, no statistical tests were conducted. All data cleaning, analysis, and figure-making was completed in R Studio software (25).

## 3.0 Results

### 3.1 Restrict Terms and Unique Award Titles

A total of 388 unique abstract titles were granted award disbursements by the NIH and CDC from 2012 to 2025 for HIV PrEP-specific research, as presented in Table 1. Of these 388 unique abstract titles, 118 (30.4%) projects would be categorized as excluded grants based on the restricted terms outlined in the methods section. Based on the award title, 47 award titles would be classified as excluded grants, while the inclusion of an award title’s abstract accounted for the majority of excluded grants at 71 award titles. Unique award titles could have restricted terms in both their title and abstract, where there were double counts and percent calculated from the total number of unique abstract titles (n = 388). For the award title, “transgender” (n =21) was the highest restricted term represented, followed by “gender” (n =14) and “LGBT” (n = 5). For an award title’s abstract, “gender” (n = 92) was a majority-represented restricted term, followed by “transgender” (n = 48) and “LGBT” (n = 24). The average number of restricted terms for unique award titles and abstracts is 1.83 restricted terms. Under a theoretical scenario, where the U.S. Presidential Executive Orders included “men who have sex with men” (MSM) within the list of restricted terms, the number of unique projects excluded would increase to 221 (56.7%) projects with the average number of restricted terms for unique award title and abstract to 2.36 restricted terms.

**Table 1.** Unique Project Grants Excluded Based on Restricted Terms. *Count is based on the aJirmative “1” input for a restricted term, where title and abstract is not exclusive and can be counted for multiple restricted terms; **Percent is calculated out of the total number of unique award titles (n = 388).

|  | <u>Count*</u> | <u>Percentage**</u> |
| --- | --- | --- |
| **Gender** |  |  |
| *Title* | 14 | 3.6% |
| *Abstract* | 92 | 23.7% |
| **Transgender** |  |  |
| *Title* | 21 | 5.4% |
| *Abstract* | 48 | 12.4% |
| **Pregnant Person** |  |  |
| *Title* | 4 | 1.0% |
| *Abstract* | 1 | 0.3% |
| **Pregnant People** |  |  |
| *Title* | 1 | 0.3% |
| *Abstract* | 1 | 0.3% |
| **LGBT** |  |  |
| *Title* | 5 | 1.3% |
| *Abstract* | 24 | 6.2% |
| **Nonbinary** |  |  |
| *Title* | 2 | 0.5% |
| *Abstract* | 3 | 0.8% |
| **Assigned Male at Birth** |  |  |
| *Title* | 0 | 0.0% |
| *Abstract* | 1 | 0.3% |
| **Average of Terms Excluded** | 1.83 |  |
| **Total Unique Projects Excluded** | 118 | 30.4% |
| **MSM** |  |  |
| *Title* | 80 | 20.6% |
| *Abstract* | 138 | 35.6% |
| **Average of Terms Excluded** | 2.36 |  |
| **Total Unique Projects Excluded** | 221 | 57.0% |

### 3.2 HIV PrEP Research Award Disbursement Characteristics

There were 2299 award disbursements across the 388 unique abstract titles for HIV PrEP-specific research from 2012-2025 from the complete grant dataset (Table 2). The NIH represented the majority of award disbursements at 2108 (92%) compared to the CDC with 191 award disbursements (8%). Scientific and health research award activities accounted for 1762 award disbursements. The recipient types varied across the award disbursements, with Junior Colleges, Colleges, & Universities comprising the most award disbursements (n =1584). There were 15 assisting listing titles represented in the award disbursements for HIV PrEP research. The highest number of award disbursements were attributed to Mental Health Research Grants (n = 1041).

**Table 2.**
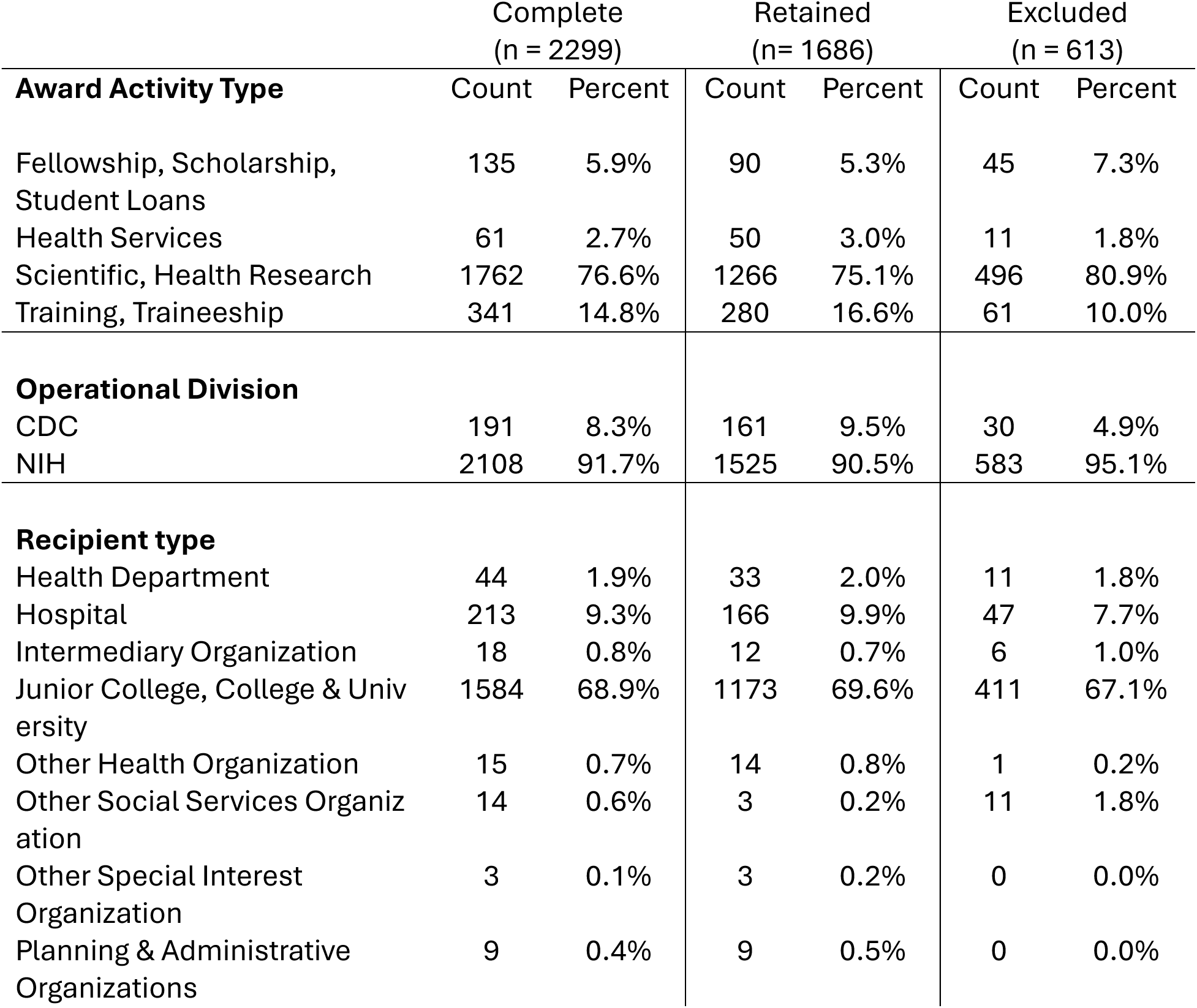
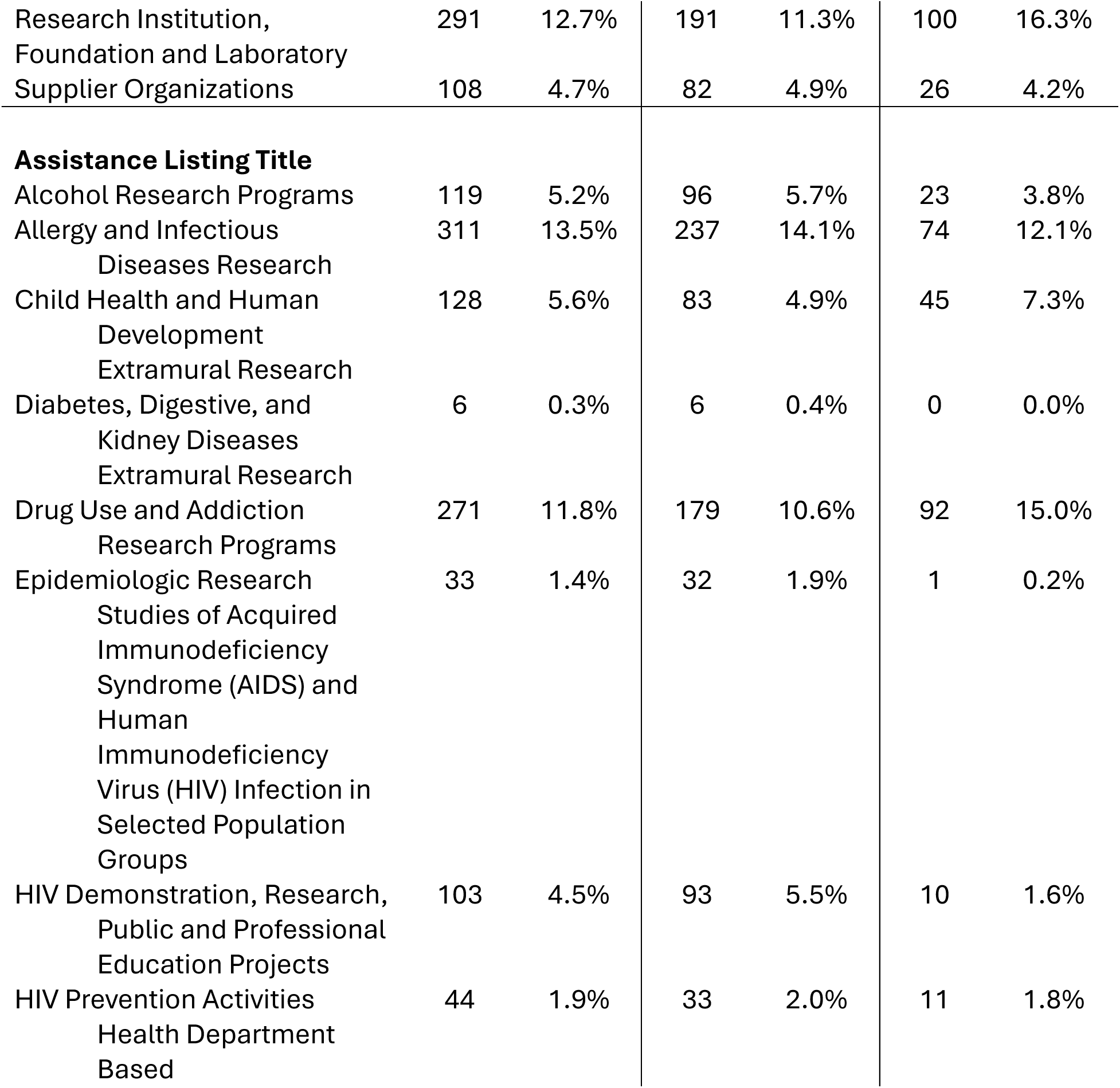
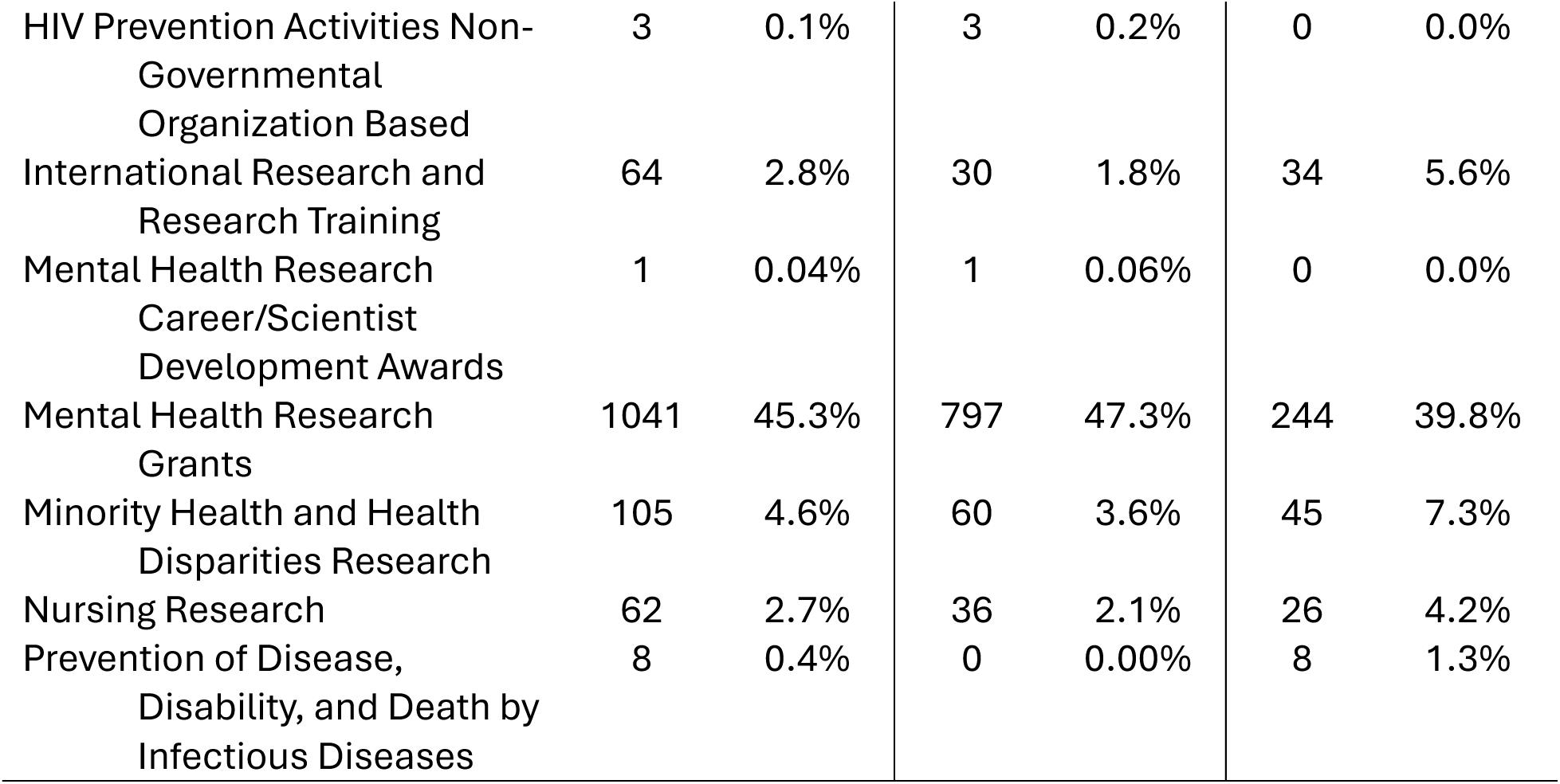
Award Disbursements Characteristics for Complete, Retained, and Excluded Categories.

When considering the excluded grant dataset award disbursements from the unique award titles, most excluded award disbursements were funded from the NIH at 95%. The scientific and health research award activity type had 496 excluded award disbursements, reflecting a greater percentage of excluded award disbursements (80.9%) than those that would be retained (75.1%). Other award activity types were similarly retained and excluded. Planning & Administrative Organizations and Other Special Interest Organizations had no excluded award disbursements, while Research Institutions, Foundations, and Laboratories had a notably greater percentage of excluded award disbursements (Table 2). The remaining recipient types were retained and excluded at similar percentages (Table 2). The majority of award disbursements excluded based on restricted terms were labeled “Mental Health Research Grants” (n =244; 39.8%), followed by the Drug Use and Addiction Research Program (n = 92; 15.0%). Three assistance listings and their award disbursements were not excluded at all: Diabetes, Digestive, and Kidney Diseases Extramural Research; HIV Prevention Activities Non-Governmental Organization Based; Mental Health Research Career/Scientist Development Awards. Only one assistance listing and its award disbursement were completely excluded: Prevention of Disease, Disability, and Death by Infectious Diseases. Based on state award disbursements, two states (Arizona and Ohio) retained zero award disbursements after applying term restrictions (only excluded) (Figure 1). When considering the retained award disbursements after applying term restrictions, five states (Indiana, Kentucky, Minnesota, Nebraska, and Oregon) had none of their award disbursements affected by the term restrictions (all retained) (Figure 1).

**Figure 1.**
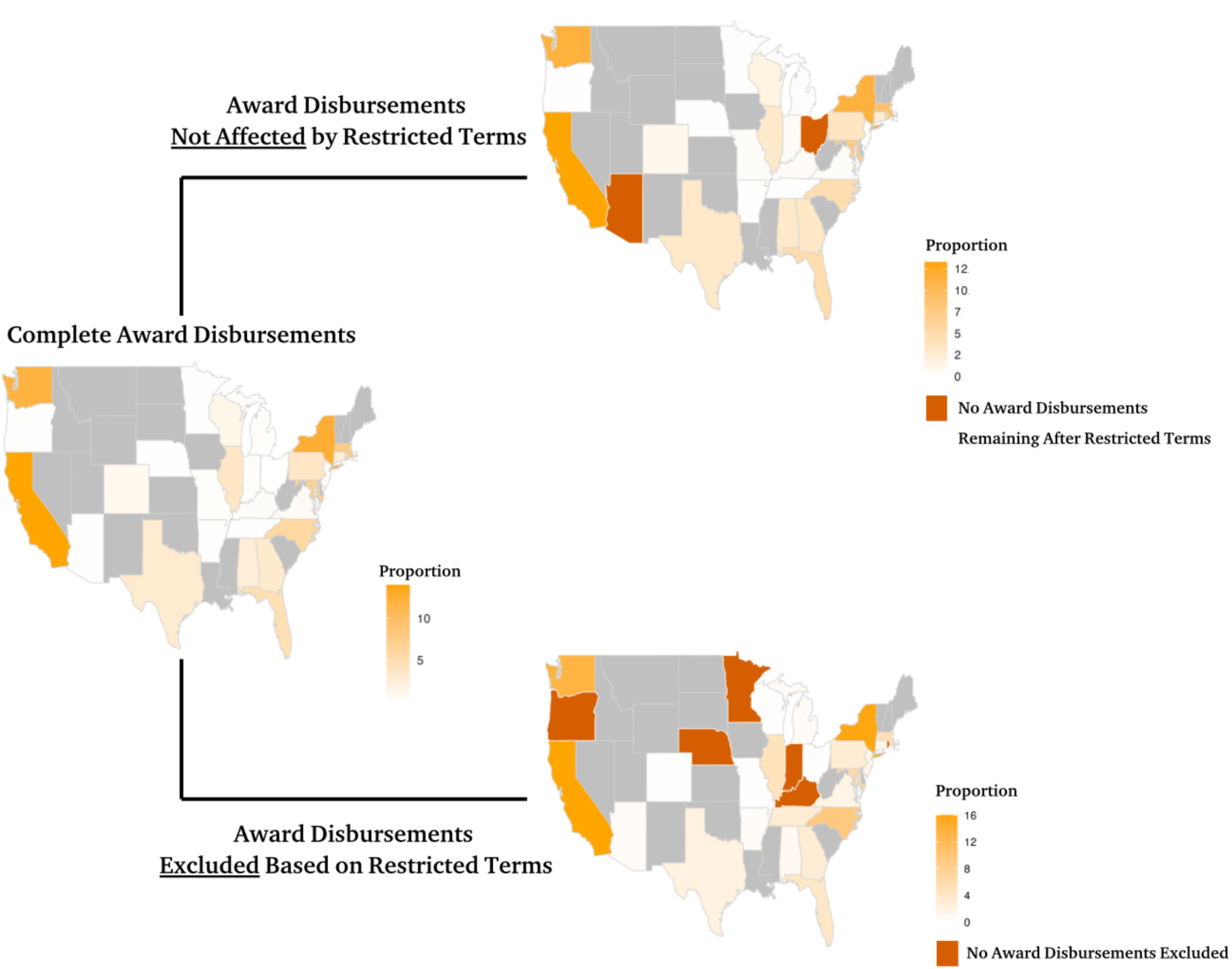
State Heat Map on the Proportion of Award Disbursements in the Complete, Retained, and Excluded Grant Datasets; Grey-filled states did not have datapoints; Burnt orange-filled states have a proportion of 0.0.

### 3.3 Elected Presidential Budgetary Period Award Disbursements

The complete grant dataset of award disbursements increased in total dollar amount for each elected presidential budgetary period since 2012 (Figure 2a). The largest increases in award disbursements were seen during the 2018-2021 elected presidential budgetary period (Figure 2a). The percentage of award disbursements for Republican states increased each elected presidential period budgetary period (Figure 2a). When assessing the retained grant dataset and its award disbursement totals, the increased trend of funding remains the same (Figure 2b). However, the rate of increase decreases from each period (Figure 2b). The percentage share of retained award disbursements increased across all periods, excluding 2012, for Republican states.

**Figure 2a.**
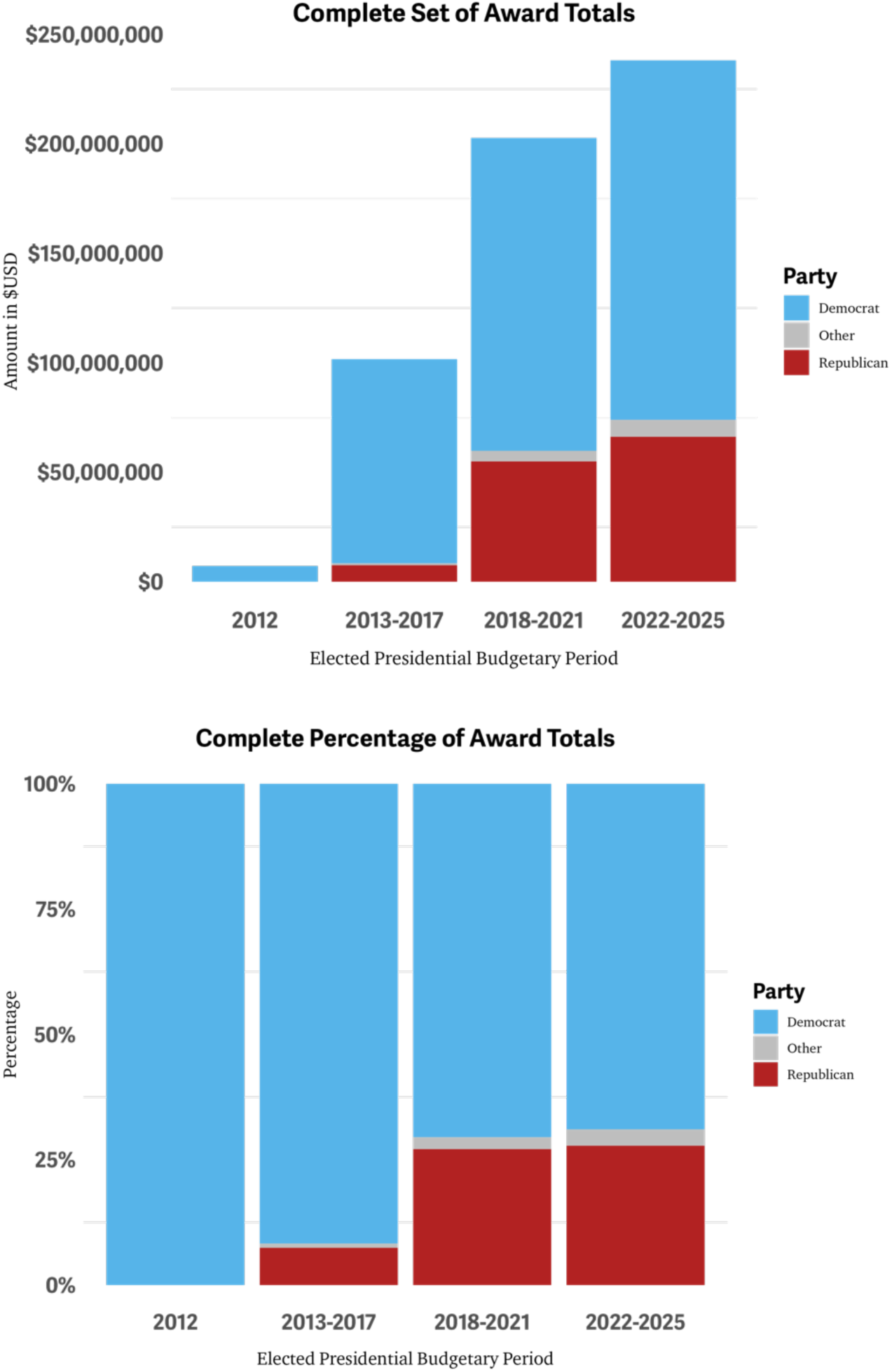
Complete grant dataset (n = 2299) of (left) total dollar amounts ($USD) and (right) the percentage of award disbursements for elected presidential budgetary periods across three political party designations (Democrat, Other, Republican).

**Figure 2b.**
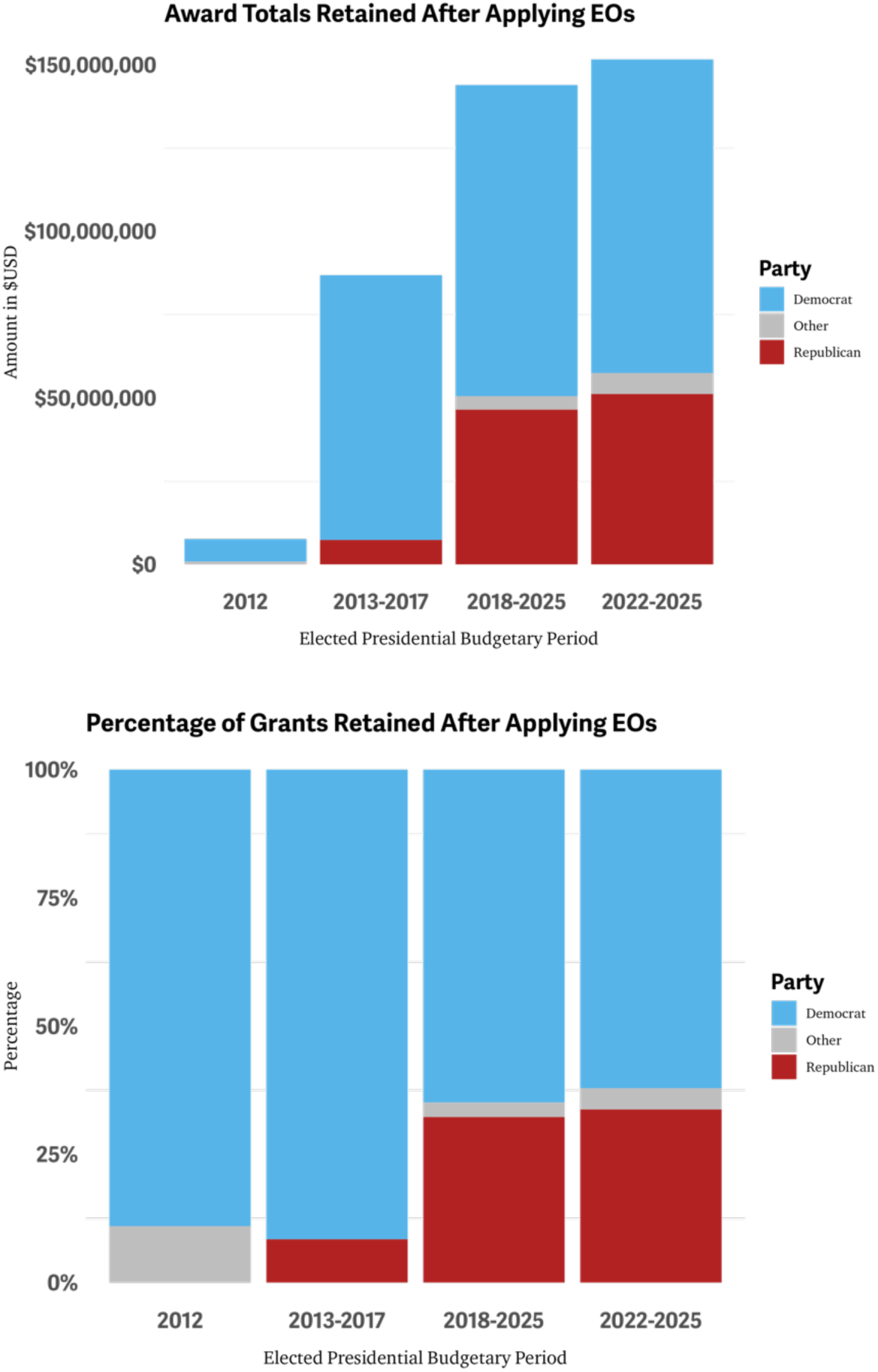
Retained grant dataset (n = 1686) of (left) total dollar amounts ($USD) and (right) the percentage of award disbursements for elected presidential budgetary periods across three political party designations (Democrat, Other, Republican).

The total dollar amount of all complete grant dataset award disbursements from 2012-2025 for HIV PrEP-specific research totaled $549,238,058 million (USD) (Table 3). The retained grant dataset showed decreased award disbursements for each elected presidential budgetary period. The difference between award disbursement totals from the complete grants minus the retain grants amounted to $158,871,814 million (USD), representing a roughly 30% decrease in total funding awarded over this time (Table 3). For the complete and retained grant datasets, the total award disbursements increased for each elected presidential budgetary period.

**Table 3.** Award Disbursements Across Elected Presidential Budgetary Periods 2012-2025. *Complete represents all award disbursements (n = 2299); **Retained represents award disbursements that were not excluded by restricted terms (n =1686).

| **Complete\*** | 2012 | 2013-2017 | 2018-20221 | 2022-2025 |
| --- | --- | --- | --- | --- |
| Democrat | \$7,196,092.00 | \$93,239,102.00 | \$142,908,724.00 | \$164,165,568.00 |
| Republican | \$0.00 | \$7,568,797.00 | \$54,583,220.00 | \$66,232,345.00 |
| Other | \$0.00 | \$834,312.00 | \$4,796,848 | \$7,713,050.00 |
| Total | \$7,196,092.00 | \$101,624,211.00 | \$202,288,792.00 | \$238,110,963 |
| **Complete Total** | | | | **\$549,238,058** |
| **Retained\*\*** | 2012 | 2013-2017 | 2018-20221 | 2022-2025 |
| Democrat | \$6,755,414.00 | \$79,530,990.00 | \$93,412,050.00 | \$94,524,819.00 |
| Republican | \$0.00 | \$7,317,045.00 | \$46,451,255.00 | \$51,168,000.00 |
| Other | \$0.00 | \$834,312.00 | \$4,067,784.00 | \$6,304,575.00 |
| Total | \$6,755,414.00 | \$87,682,347.00 | \$143,931,089.00 | \$151,997,394.00 |
| **Retained Total** | | | | **\$390,366,244** |
| **Difference by Period** | **-\$440,678** | **-\$13,959,864** | **-\$58,357,703** | **-\$86,113,569** |
| **Difference between Complete minus Retained Totals** | | | | **-\$158,871,814** |

## 4.0 Discussion

HIV PrEP represents the most significant modern-day biomedical advancement in the 21st century thus far in ending the HIV epidemic. Specific research regarding HIV PrEP advances many fronts, including biomedical innovation, population and demographic receptiveness, uptake, implementation, and so on. Additionally, funding for these research endeavors supports the next generation of researchers, which ultimately sustains progress in ending the HIV epidemic in the U.S. and beyond, along with maintaining robust public health response. A funding loss of $158,871,814 from 118 unique award titles since 2012 based on restricted terms around gender and sexuality exemplifies a devastating reality of what the current condition means for the future of HIV PrEP research. This research is the first to retrospectively characterize how EO 14168 implementation would impact a subset of HIV research on HIV PrEP.

### 4.1 Clear Targeting of Gender & Transgender Representation

Restricted terms “gender” and “transgender” from EO 14168 were primarily targeted for exclusion, citing 175 instances of using either term across grant titles and abstracts. Gender is a central social determinant of health as health outcomes are shaped through differential exposure to intermediary determinants of health (26). Nearly one-fifth of new HIV diagnoses in 2022 were among women (27), where Black/African American females account for 50% of the new diagnoses amongst people assigned female sex at birth (28). In 2024, the NIH Office of AIDS Research and Office of Research on Women’s Health set forth an ambitious aim to address HIV amongst women and gender-diverse people. Central researchers from the NIH stated, “The time is now to center the health of women, girls, and gender diverse people across the HIV research continuum” (29). According to data in 2022, only 15% of females who could benefit from HIV PrEP were prescribed it, compared to 40% of males who could help (30). Undoubtedly, a restriction on the term “gender” would be consequential in influencing the directive of research regarding women in HIV and ultimately stifle innovative research in HIV. Transgender and gender-diverse (TGD) populations are often not represented in HIV research and not engaged in clinic care or trials (31). Despite this, transgender women have more than 48 times the odds of HIV infection compared to the general population (32). Gianella et al. argue that culturally appropriate research opportunities advance science while also empowering TGD populations to combat this disparity (33). A systematic review of HIV PrEP uptake among transgender populations found lower PrEP uptake based on a lack of trust in medical institutions and the influence of social networks (34). Erasure by restricting “transgender” in research grants undermines progress in the healthcare of TGD in the U.S. and around the world. In either case, women, transgender, and gender-diverse people have persisted and resisted since the beginning of the epidemic to advocate for a better HIV response and resist attempts to subvert the importance of their existence in society (35,36). Gender-diverse and transgender people have an enduring history since early civilization, suggesting their persistence and resilience in society will remain despite hostile political climates (37). For instance, transgender women living with HIV operate helping networks to maintain and improve the health of their communities despite continued opposition and discrimination from the healthcare system and society in the form of stigmatization, access to culturally competent services, and lack of medical knowledge to provide trans-care (38). Eliminating “transgender” in research grants and award disbursements is transphobic, where “institutional homophobia/transphobia includes the ways that institutions discriminate against people based on their sexual orientation or gender” (39). Ultimately, the compromise of research by EO 14168 implementation for TGD and women in HIV will exacerbate HIV-related disparities, allowing for the HIV epidemic to persist.

If “men who have sex with men” or MSM was added as a restricted term under a theoretical scenario, more grants would be excluded (n = 221). The uncertain nature of selecting restricted terms could destabilize entire research areas for specific populations if presidential executive orders broadened their scope. The broader restrictions would indicate that nearly 60% of previously conducted HIV PrEP-specific research since 2012 would have been restricted, severely hindering advancements in ending the HIV epidemic.

### 4.2 Excluded Award Titles were Diverse and Crucial Research Endeavors

Training the next generation of scientific researchers is essential if the U.S. and the globe plan to end the HIV epidemic (40). While composing a smaller percentage of excluded grants (17.3%), training and related funding are crucial to sustaining the continuity of research endeavors, which drives knowledge advancement and application, human capital development, and the federal government’s mission (41). Since the beginning of the HIV epidemic, U.S.-based researchers have been at the forefront of primary innovation, including the first treatment for HIV through clinical trials (42), the first human trial of HIV PrEP (43), and the success of long-acting injectable treatment and prevention modalities (44,45). The U.S. federal government, primarily the NIH, funded these research and clinical trials. Undue restrictions based on key terms relevant to research and its reproductions for supporting the training of research would stifle the U.S.’s presence as a research frontier for HIV. These restricted terms also impacted the research institutions that house these trainees. Rhetorical tactics to decontextualize the work of colleges and universities by Republican and Trump entities undermine the contributions of these institutions to research and the economy (46,47). This data shows how excluding grants negatively impacts the funding received in states that have voted Republican in elections since 2012. Colleges and universities in these Republican (”red”) states recently pushed back to funding cuts by the NIH (48). Colleges and universities are not the only institutions impacted by these restricted terms. Hospitals account for nearly 8% of the award disbursements excluded. Based on Trump’s and the Republican party’s agenda, hospitals may sustain more loss, impacting the ability to conduct research and provide care to patients (49). Additionally, supplier organizations would have excluded their award disbursements. EO 14168 would limit the continuum of research from development to implementation. Supply chain organizations are crucial to sustaining healthcare delivery and preserving the health of Americans (50). The blanket exclusion of grants is not selectively targeting one type of institution but compounding the negative impacts by impacting all facets of the research process regarding HIV PrEP research.

A notable finding from this research is the number of Mental Health Research Grant award disbursements that would have been excluded (n = 244; 39.8%). Existing research has pointed out how, despite the advancements in biomedical technologies, mental health is a key factor in HIV prevention and transmission (51). Mental health is a clearly cited factor in the relationship of PrEP uptake, especially for those experiencing depression (52). There is active discussion on how prescribing PrEP in community mental health settings can better align with the needs of those who could benefit from PrEP through the integration of sexual and mental health services (53). Furthermore, some researchers argue that the integration of PrEP and mental health services increases HIV PrEP’s effects and scale-up efforts for uptake and implementation (54). Neglecting the findings of these data stalls the full realization of biomedical advancements like HIV PrEP from impacting the HIV epidemic in the short and long term. Hence, mental health grants that are excluded based on these restricted terms hold the relevant results to better health outcomes and reduced HIV transmission. People vulnerable to acquiring HIV are also impacted by substance use problems that heighten the effects of barriers to accessing the HIV prevention landscape, including social and economic factors (51). Award disbursements for drug use and addiction programs were excluded at a higher percentage (15.0%) than those retained (10.6%). Political differences between Democratic and Republican party affiliations are key factors when understanding public opinion and response to the opioid and substance use crisis (55). While many Republicans have recently opposed research and tools against substance use and addiction programs (56), constituents in their districts are more likely to be impacted by cutting these services, especially in rural areas (57). Researchers suggest the integrated service models for HIV PrEP and harm reduction offer an optimal setting to increase access and support better health outcomes (58,59). However, ongoing research is necessary to establish these services’ feasibility and broader impact, where HIV PrEP-specific research plays a crucial role in evidence generation. Research is not sufficient to deliver harm reduction services, but it is a vital component (60). The intersectional nature of mental health and substance use in HIV prevention is underscored in the literature (61,62). Stalling research on this front will diminish the valuable returns of new and emerging strategies to tackle this front of the HIV epidemic in the U.S.

### 4.3 Potential Research Funding Loss is Non-Negligible

Most of the excluded award disbursements came from the NIH (n = 583; 95.1%). Direct economic contributions from NIH funding estimate that for every $1 (USD) spent, approximately $2.46 of economic activity is generated (63). Applied to the findings of this research with the caveat of a small percentage associated with CDC funding, a total exclusion of $158,871,814.00 for HIV PrEP-specific research would have seen a roughly $391 million dollar decrease in economic activity from 2012-2025, not adjusting for inflation. This loss in investment and economic activity is non-negligible. Investment in research and development on the innovation frontier can protect a country’s growth during an economic crisis, like a recession (64). As a leading pioneer in HIV PrEP advancements, the United States would face a great amount to lose economically and intellectually through these restricted terms.

### 4.4 Both Democrat and Republic States were Impacted to Varying Degrees

The loss of award disbursements for HIV PrEP-specific research negatively impacted both states that voted Democrat and Republican across the presidential election periods. Interestingly, Indiana, Kentucky, and Nebraska saw none of their award disbursements excluded by the restrictions, all of which cast electoral votes for the Republican party. Minnesota and Oregon were the Democrat states with no award disbursements affected. Notably, Arizona has consistently leaned Republican since 2012, and Ohio, with recent support for the Republican presidential candidate, saw all of their award disbursements excluded based on the restricted terms. Republican states were more negatively impacted by the Executive Order than Democratic states for total exclusion of grants related to HIV PrEP. However, it is worth noting that Republican states retained a higher percentage of award disbursements when excluding grants, presenting a possible preference for these executive orders to “red” states for HIV PrEP research.

Republican lawmakers, since the beginning of the current administration, have carved out exemptions for their constraints for spending cuts (65). While these executive orders do indeed target gender, sexual orientation, and gender expression, they also appear to target states that lean Democrat for presential elections, expanding the impact of these executive orders beyond ideology and into political party differences. In either case, while the economic fallout for each Democrat and Republican state would be experienced differently, the data indicates economic activity for a majority of states would be readily impacted.

### 4.5 Research Strengths

The primary strength of this research is the quantified loss of funding and the extent to which the EO on gender and ideology would have impacted HIV PrEP-specific research from 2012-2025. Additionally, the breakdown of what type of institutions are impacted, along with a state-level analysis with political affiliation, offers a nuanced understanding of the research impact of the current administration, which is paving for the future. The framework of this research endeavor can provide a reasonable basis for understanding how other research areas would be impacted.

### 4.6 Research Limitations & Future Directions

This research is not without its limitations. Importantly, this research narrowed its scope by selecting award disbursements from only the award title and included “PrEP” or “pre-exposure prophylaxis,” narrowing the number of total grants related to HIV PrEP research that did not contain these term criteria in the title. Any missing award disbursement amounts from TAGGS may skew funding differences, impacting the results’ magnitude. The retrospective nature of this research is merely hypothetical of what would have occurred if these EOs were applied from 2012 to 2025, which does not articulate the current or future restrictions on research. The small award disbursement counts across the unique grants meant that statistical testing could not find significant differences between various variables and groups. EO 14168 on gender ideology was the source of our restricted sources. However, EO 14151, which aims to end diversity, equity, and inclusion initiatives, provides a longer list of prohibited or flagged terms for implementation (66,67), which may have drastically changed the results of this data.

Future research will benefit from more robust models that can act in a predictive manner to understand political shocks and shifts in HIV PrEP research. Additionally, more research is required to value the entire scope of HIV research, including prevention, care, and treatment.

## 5.0 Conclusion

HIV PrEP-specific research comprises only one piece of the large pie of HIV research, where a loss of nearly $160 million dollars is non-negligible and likely to have impacted the response to the HIV epidemic in a consequential way since 2012. Appropriation committees in the U.S. House of Representatives and Senate should consider this retrospective analysis when considering budgetary cuts, including the extensive negative economic impact on their constituents. The way forward to ending the HIV epidemic should not half-hazard target grants and award disbursements based on ideology through restricted terms but rely on evidence and constituent voices to make clear decisions on the budget. If a critical part of a mechanic’s toolbox, such as a wrench, is reduced, how can we expect this mechanic to work with what they have and fix the car in front of them? HIV PrEP research is our wrench in working towards ending the HIV epidemic.

## Data Availability

The availability of this data is available upon request via email with the main author.

